# A realist interview study of a participatory public mental health project “#KindnessByPost”

**DOI:** 10.1101/2022.12.17.22283620

**Authors:** Hannah Rachel Scott, Katey Warran, Kathleen Fraser, Beverley Chipp, Gail McGinnes, Mike Towers, Brynmor Lloyd-Evans, Luke Sheridan Rains

## Abstract

**Background:** #KindnessByPost (KbP) is a participatory public health initiative in which people anonymously send and receive cards containing messages of goodwill with others also taking part in the programme. Quantitative evaluations of KbP consistently find evidence of improvements to people’s mental wellbeing and feelings of loneliness after participation and three months later. Our aim in the present study is to develop a programme theory of KbP, which describes for whom the KbP intervention improves mental wellbeing, other reported impacts, in which contexts it has these effects, and the mechanisms by which it works

**Methods:** We use a realist interviewing methodology to develop the programme theory. We conducted a focus group with the KbP executive team, and 20 one-to-one interviews with KbP participants. During analysis, a co-production working group iteratively developed a Theory of Change model comprising context-mechanism-outcome statements [CMOs] to map out the mechanisms present in KbP.

**Results:** We developed 145 CMO statements, which we condensed and categorized into 32 overarching CMOs across nine thematic topics: access to scheme; pathways to involvement; resources; culture; giving post; receiving post; content of received post; community; long term impact. These CMOs set out pathways through which KbP benefited participants, including from doing something kind for someone else, of receiving post and appreciating the effort that went into it, and from the creative process of creating post and writing the messages inside them. Effects were sustained in part through people keeping the cards and through the social media communities that emerged around KbP.

**Discussion:** Both giving and receiving post and the sense of community benefited participants and improved their mood and feelings of connectedness with others. Connection with a stranger, rather than friends and family, was also an important feature of the initiative for participants. Our wide range of CMO pathways by which KbP produced positive outcomes may mean that the intervention is applicable or adaptable across many communities and settings. Taken together with evidence from the quantitative evaluations, KbP is potentially an effective, low-cost, and highly scalable public health intervention for reducing loneliness and improving wellbeing.

## Introduction

Interventions designed to improve wellbeing are often resource-intensive, leading to high and often prohibitive costs for health providers and limited access for potential recipients. Participatory public health interventions – programmes that are primarily powered by communities themselves – have the potential to relieve burden on healthcare providers by offering low-intensity support for health issues that present a low clinical risk [1] and can have a preventative role by maintaining wellness and social connection. With participatory interventions, communities are afforded a sense of ownership over a support system and can shape it in a way that meets their needs; they can also be run at a low cost with minimal need for clinician input if delivery is primarily facilitated by the community. Additionally, in cases where participants both deliver and use the intervention, typical structural power imbalances between service deliverer and service user roles are not present, potentially making the intervention more accessible [2]. With a clear understanding of the outcomes that these interventions achieve, and the mechanisms through which they do so, participatory public health interventions have the capability to be replicated across communities and scaled up to the population level, offering practical solutions to improving, or at least protecting, public health.

#KindnessByPost (KbP) is a participatory public health project in which people send and receive cards or letters containing messages of goodwill. It is conducted through regular card exchanges organised by the non-profit organisation Mental Health Collective [3]: Anyone living in the United Kingdom (UK) and over 13 years old, or younger if in partnership with an adult, can take part. People sign up for an exchange via a dedicated website [4]. They send a card to one randomly allocated person and receive a card from a different person also participating in the exchange. They are given only the first name and the address of the person they are sending a card to. If someone does not receive a card at first, they can use a ‘back-up system,’ in which participants volunteer to send extra cards to avoid people missing out. The Mental Health Collective estimates that 97% of people receive a card [4], and people are informed that if they do miss out, it is not personal since the exchange is anonymous. Several KbP exchanges take place each year; and as of 9th December 2022, there have been 21,304 unique registrations in total for people taking part [5].

KbP has the potential to benefit participants in numerous ways: through both the act of doing a kind act and the receipt of kindness; through engagement with the creative act of making a card or writing a message; and through the sense of community or time spent together exchanges often create, such as parents engaging their children or teachers engaging their classes in working on a card together, or the use of social media by people to post and comment on the cards they receive using hashtags. There is an established relationship between improved mental health, and engagement in leisure activities and their associated communities [6]; the mechanisms through which this association happens are complex and act at multiple levels [7]. Arts-based interventions are also supported by a wealth of evidence [8], with studies suggesting that positive impact can be through using art as a way to develop social connections as well as the act of creating and expressing oneself being an opportunity for stress-relief and empowerment [9, 10]. Studies of carrying out simple, kind acts towards others (Dunn et al., 2008) and being part of a group exchange of kind acts [11] have also found a positive impact on wellbeing.

An evaluation of the KbP 2020 #GreatBritishValentine event [12] found significant improvements in people’s feelings of wellbeing, loneliness, sense of belonging, and hope. Qualitative analysis of participant feedback was overwhelmingly positive. However, this evaluation had a relatively low response rate, only followed up participants immediately after the card exchange, and was only able to offer broad insights into potential mechanisms through which it brought about positive effects. More extensive quantitative evaluation of the interventions has since taken place, showing that participation was associated with a significant increase in feelings of wellbeing and a decrease in feelings of loneliness; that these improvements were sustained at 3-months follow-up, and that the estimated organisational costs of the programme were very modest (<£1 per participant) [13].

We sought to use qualitative methods to evaluate KbP: a participatory public health programme. An area where understanding is still relatively lacking is in understanding the mechanisms by which interventions such as these produce positive outcomes [7]. Qualitative evaluation of the intervention allowed us to examine not only the intended outcomes of the intervention, but also how participants interact with a kindness intervention with a creative and community-based focus to achieve these outcomes. Specifically, we took a realist approach [14] to data collection and analysis, aiming to develop an in-depth understanding of the initiative’s benefits, mechanisms, and contextual factors affecting its delivery and experience. This will explore the extent to which the KbP model resonates with a range of communities and the flexibility of its application. A richer understanding of this promising participatory public health programme will help understand how and in what circumstances a reciprocal message of kindness received by post can contribute to the wellbeing of a person with mental health challenges.

Our aim in the present study is to use qualitative methods to develop a programme theory for KbP and explore for whom the KbP intervention addresses loneliness and improves wellbeing, in which contexts it has this effect, and the mechanisms by which it works.

## Methods

### Study design and Setting

We adopted a realist interviewing approach [14, 15] a method recommended in the Magenta Book [16] for developing and refining programme theories of social and healthcare interventions. It is a theory-based approach that seeks to understand ‘what works, for whom, and in what circumstances?’

Data collection and analysis followed the process outlined by Manzano [17], where a programme theory is developed through distinct phases of realist interviewing: 1) theory gleaning interviews to establish initial theories of how the intervention works as perceived by intervention designers; 2) theory refinement interviews to modify and clarify theories; 3) theory consolidation interviews to strengthen support for theories and establish a logical understanding of how, why and in what circumstances the intervention works. In line with this, recruitment and analysis were iterative processes conducted in conjunction with each other.

Data collection and analysis were conducted iteratively, with multiple cycles of participant recruitment and interviewing followed by a phase of analysis, guided by regular meetings of a co-production working group. The working group consisted of four academic researchers and four lived-experience researchers with experience of participating in KbP exchanges and, for some, of experiencing mental health problems and loneliness. Working group members were given training in realist methods where needed prior to data collection starting. This group steered data collection, conducted the data analysis, and iteratively developed a programme theory of KbP.

We conducted this work in the context of two KbP exchanges in Spring and Summer 2021. KbP exchanges are marketed via newspapers, radio, Mental Health Collective mailing lists, social media, and word of mouth. People sign up for a KbP exchange via a dedicated website [4].

### Focus group

Data collection began with a focus group held with two members of the KbP executive team who developed and run the KbP initiative. It followed a semi-structured interview schedule, developed by HRS, LSR and BLE, to draw out their perspective of why and how the programme impacted its participant s. The group was conducted using video conferencing software and audio recorded. The recording was then anonymised and transcribed for analysis. From this data, an initial Theory of Change model was developed by HRS, LSR, BLE, and KW that was expanded and refined during subsequent data collection and analysis stages.

### Individual interviews and working group meetings

Following the 90-minute focus group, one-to-one interviews lasting up to 60 minutes were held with KbP exchange participants. Recruitment and interviewing occurred in cycles of four to five participants at a time, followed by a meeting of the working group, who would analyse the interview data from the previous set of participants and make subsequent decisions about revisions to the interview topic guide and the next phase of data collection.

KbP participants were able to register interest to participate in the present study and consent to their information being used for recruitment when completing a questionnaire conducted for a concurrent evaluation conducted by the same research team [13]. We recruited to the present study purposively, aiming to generate a demographically diverse sample of participants with a range of different experiences of KbP, including participants from different ethnic backgrounds, genders and ages, and those who had had diverse levels of participation in KbP (e.g. first time vs repeat participants, those who had missed receiving a card in the exchange). A KbP staff member invited purposively identified individuals via email to contact the interviewing researcher (HRS). HRS then arranged an initial phone conversation to explain the study fully to the participant and arrange an interview, held at least 24 hours later. Consent was obtained at the beginning of the interview. Interviews were conducted by telephone or video conferencing software according to participant preference. They were audio recorded and transcribed prior to analysis. At the beginning of each interview, the interviewer (HRS) briefly explained the concept of realist interviewing to the participant.

An initial interview schedule was developed and reviewed with the working group prior to the first set of participant interviews, covering reasons for participation, the experiences of giving and receiving post, accessibility, and recommendations for improvement. At each working group meeting, the interview schedule was reviewed by the group and revised as needed based on analysis discussions. For example, questions may be added or revised to further explore specific topics, or to refine or refute emerging theories about the intervention; questions were removed if they were unlikely to add any further value to the analysis. Over the course of the process, questions moved from being exploratory (theory gleaning), to refining and consolidating the working group’s theories, which would often be discussed with participants during interviews, alongside the interview schedule questions.

### Analysis

Data analysis sessions were alternated with phases of participant interviews to enable analysis discussions to guide future data collection. The data analysis was performed by the working group through five meetings held approximately every two weeks: four times following each cycle of participant interviews, and once to finalise the results of the data analysis. For the first four meetings: before the meeting, members reviewed transcripts; during the meeting, members reviewed interview transcripts, developed theories about the intervention, and developed/revised the KbP programme theory, depicted in a Theory of Change model.

Context-mechanism-outcome configurations [CMOs] were used as analytical tools to construct and express theories about the KbP intervention. CMOs represent a way to set out a causal relationship between the context in which an intervention operates, the mechanisms that operate in this context, and the outcomes that the mechanisms produce [15]. Researchers analysed interview transcripts to extract explicit and implicit CMOs. Working group members would then discuss proposed individual contexts, mechanisms, and outcomes as well as the complete CMOs; and suggest new ones. The working group created a Theory of Change model to visually represent the CMOs, including the variations in outcomes for participants through different contexts and mechanisms. Members would discuss changes or additions that should be made to the Theory of Change model as theories became more refined through data collection.

Once all the data had been collected, authors HRS, LSR, BLE, KW carried out an analysis of all the transcripts to confirm that the CMOs in the Theory of Change model were firmly grounded in the data, and to check for any CMOs that were missing from the model. This resulted in an extended list of 192 CMOs, which HRS condensed into 145 statements by combining or deduplicating CMOs with very similar content, and these were reviewed by LSR. These were then summarised by HRS, LSR, BLE, and KW in 32 overarching CMO statements and categorised into thematic topics.

During the final working group meeting, the list of 32 overarching CMO statements was reviewed by all working group members, refining, and clarifying language. Following this, authors HRS, LSR, BLE, KW used the CMO list and supporting statements to write summary text to accompany each set of topic- focused over-arching CMOs, which are presented in the results section.

## Results

We conducted interviews between May and October 2021. Two members of the KbP executive participated in a single focus group (one male, one female; both White British). Twenty participants took part in one-to-one interviews. Of the interview participants, fifteen identified as female, five identified as male; fifteen identified as White British, five as Asian/Asian British. Participants were aged between 18 and 72, with a mean age of 38.

### Context-Mechanism-Outcome Statements

In total, 145 CMO statements were identified, which are presented in Appendix 1. For ease of understanding, these were categorized into nine thematic topics representing distinct aspects of the exchange process, and summarised into 32 overarching CMO statements, which are presented below:

#### 1. Participation > Access to scheme

Participants often felt that the scheme was appealing because it offered a manageable and structured way to be kind to somebody without having to maintain an ongoing relationship (table 1); often contrasting it to having a pen pal, which would require more time investment and did not feel feasible to many. Similarly, people valued the relative anonymity and security of only needing to provide a first name and postal address for participation. The timing of exchanges was also important; participants liked not having to commit to more than one exchange at a time and liked that they were infrequent enough not to feel routine and something that ought to be done. Some participants also felt that having a theme for each exchange helped them to engage with it and could be helpful in giving them something to focus their post on.

**Table 1:** Access to scheme CMOs

| Overarching CMO statement | Illustrative participant quotation |
| --- | --- |
| 1. If people can choose which/how many of the several exchanges each year they participate in, and they only need to commit to producing one card each time, then they are more likely to participate in an exchange, because doing so feels manageable and flexible enough for them to fit into their lives when/if they can. | *"I have no capacity for a pen pal, that doesn't interest me. I like the idea of sort of love bombing someone from a distance and then to possibly getting my card in return, that's enough for me."*<br>P17 |
| 2. If there is a broad theme to the exchange then people may be more likely to participate because it gives people a framework for what to create and something they can emotionally connect with. | *"I try and work around the theme in terms of decorating the card to go alongside it. If I can't do that then I'll try and put something inside to reflect it that way."* P19 |
| 3. If people see diverse kinds of post on social media, rather than only high value or highly creative posts, then they may be more likely to participate, because they do not think they need | *"Some of them are just beyond amazing - these people really go to quite lengths to make these cards. So that can be pretty intimidating, because then I'm like, oh my God I don't have time, I can't* |
| certain skills or need to meet a certain standard to participate. | *paint like that, just I can't do that. But then I just have to say, well what I send out is what I send out and I hope that it makes a difference."* P4 |
| 4. If people only have to provide a first name and address for the exchange, then they may be more likely to participate because the anonymity of this feels reassuring and limits potential negative outcomes. | *"Nowadays people are worried about their details and stuff, but this was like it's good that it's anonymous and that the person you're sending the card to is different from the person that you're receiving it from.... Yes, so I didn't have to give away my personal details."* P9 |
| 5. If the online platform/website that facilitates exchanges is user-friendly then people are more likely to participate because it is a quick and straightforward process for people to sign up, and guidance on the website answers any questions they have. | *"A few little ground rules were useful so there was a sense of expectation to fit into that was really not very narrow or restrictive, but it just set those sort of parameters that was quite helpful and reassuring."* P7 |

Participants who used social media noted that it could have both a positive and negative influence on their choice to participate. Often, it served as inspiration for peoples’ own cards. However, for some, seeing particularly creative cards or post that included gifts could be off-putting if they did not see themselves as sufficiently creative or did not think that sending gifts was in the spirit of the exchange. Some participants thought that it was important to see simple creations and cards, thereby communicating a message that you do not need to ‘be creative’ or artistic to take part.

A small number of participants commented on the KbP website, saying that it was user-friendly and that the guidance for cards provided useful advice.

#### 2. Participation > Pathways to involvement

Most participants became involved in KbP after hearing about the initiative from somebody they knew (table 2); most frequently this was because a contact had shared something about it on social media. This personal endorsement made participants feel more comfortable trusting the exchange as they knew somebody who had had a positive experience with it. A number of participants spoke about how they themselves shared their experience of participation online or talked to others about the project to help promote it.

**Table 2:**
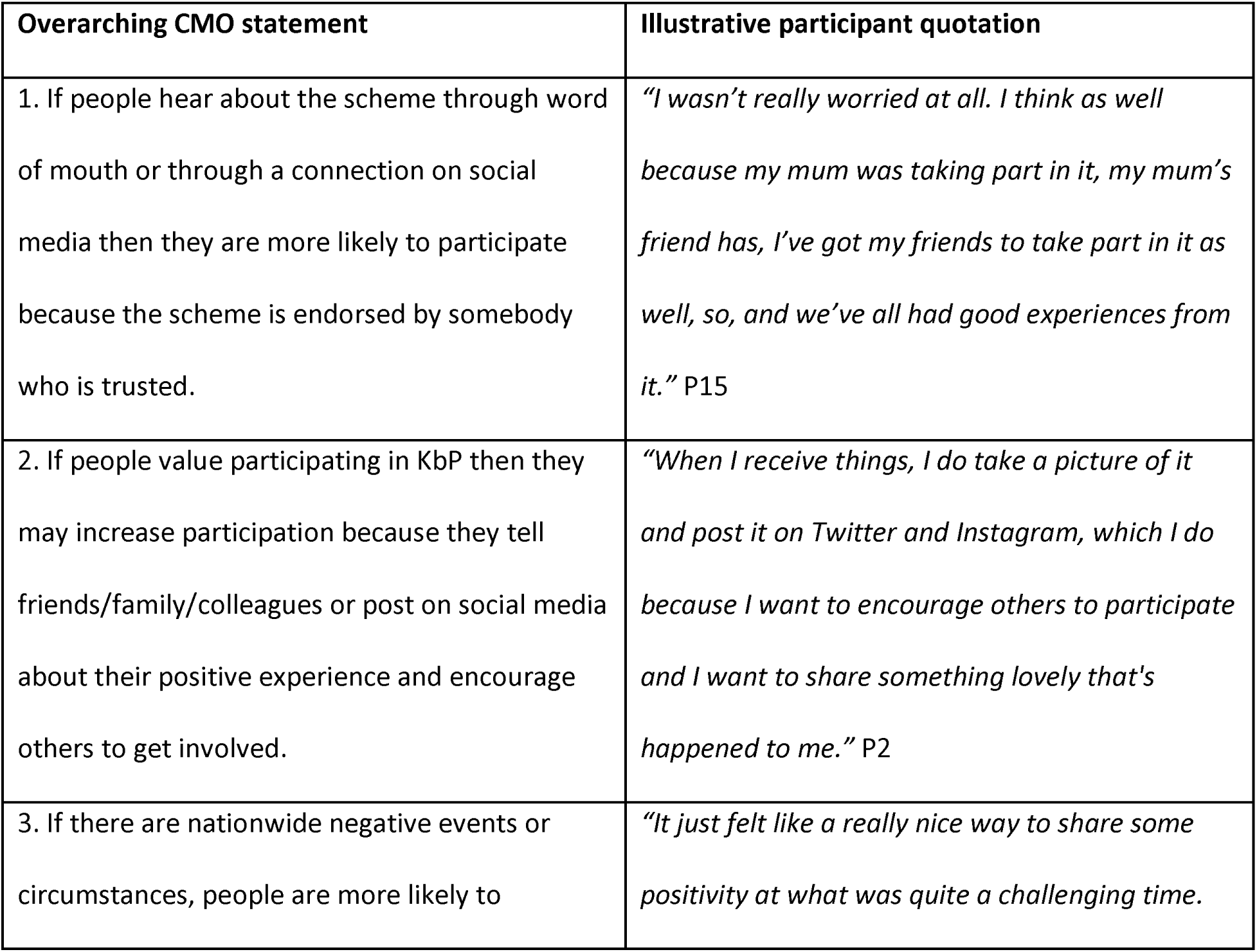
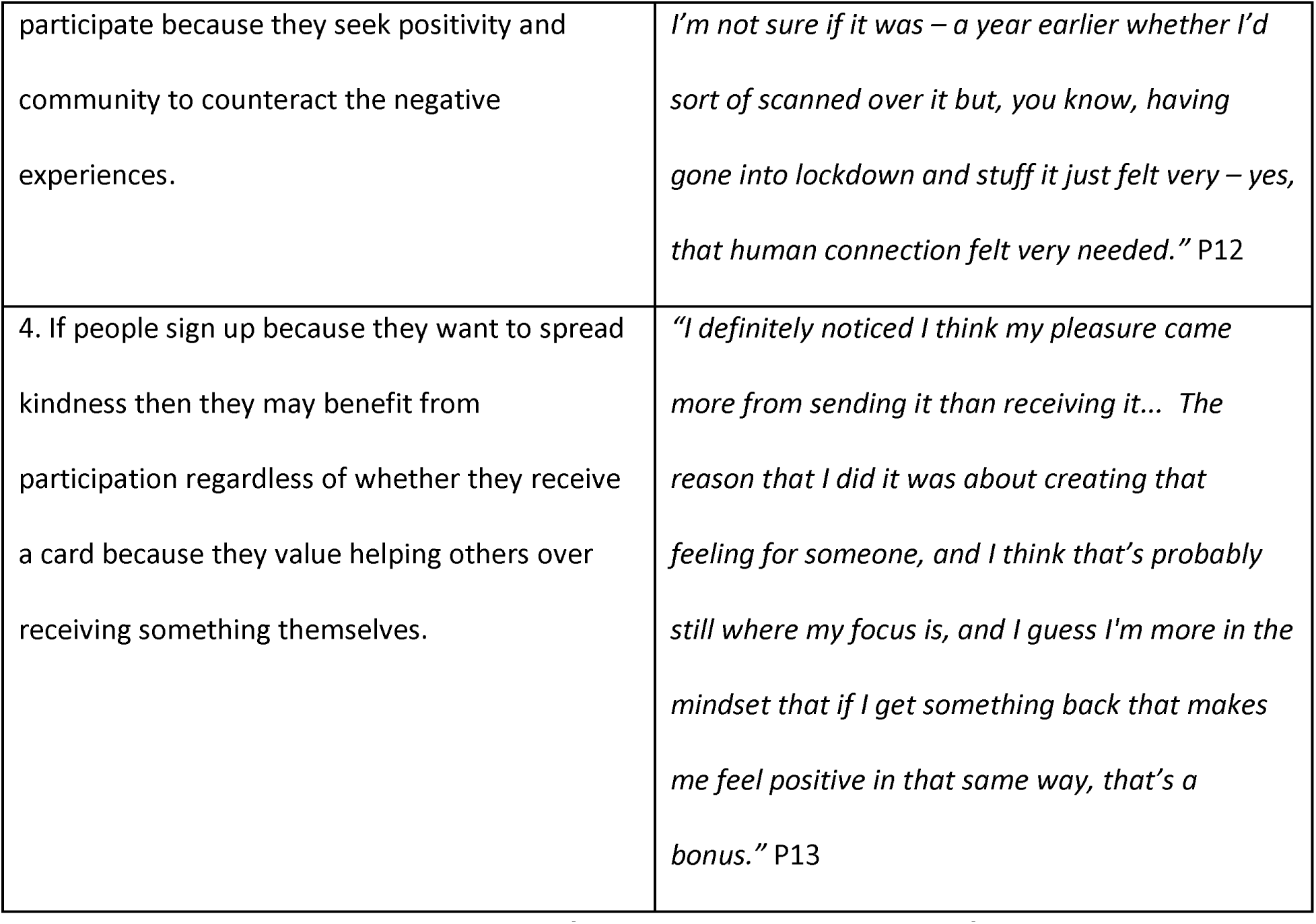
Pathways to involvement CMOs

| Overarching CMO statement | Illustrative participant quotation |
| --- | --- |
| 1. If people hear about the scheme through word of mouth or through a connection on social media then they are more likely to participate because the scheme is endorsed by somebody who is trusted. | *"I wasn't really worried at all. I think as well because my mum was taking part in it, my mum's friend has, I've got my friends to take part in it as well, so, and we've all had good experiences from it."* P15 |
| 2. If people value participating in KbP then they may increase participation because they tell friends/family/colleagues or post on social media about their positive experience and encourage others to get involved. | *"When I receive things, I do take a picture of it and post it on Twitter and Instagram, which I do because I want to encourage others to participate and I want to share something lovely that's happened to me."* P2 |
| 3. If there are nationwide negative events or circumstances, people are more likely to | *"It just felt like a really nice way to share some positivity at what was quite a challenging time."* |
| participate because they seek positivity and community to counteract the negative experiences. | *I'm not sure if it was – a year earlier whether I'd sort of scanned over it but, you know, having gone into lockdown and stuff it just felt very – yes, that human connection felt very needed.” P12* |
| 4. If people sign up because they want to spread kindness then they may benefit from participation regardless of whether they receive a card because they value helping others over receiving something themselves. | *“I definitely noticed I think my pleasure came more from sending it than receiving it... The reason that I did it was about creating that feeling for someone, and I think that's probably still where my focus is, and I guess I'm more in the mindset that if I get something back that makes me feel positive in that same way, that's a bonus.” P13* |

Participants tended to sign up because they wanted to be kind to others, rather than primarily hoping to receive post themselves. Some were particularly motivated by the COVID-19 pandemic and wanted to seek connection and positivity during a challenging and isolating time. A small number of participants did exchanges with their children, wanting to facilitate or encourage their prosocial behaviour.

#### 3. Resources

Some participants commented on practical barriers to participation (table 3). This could be a lack of financial resources to buy the materials needed to send a card, or a lack of time in which to participate. Some participants did, however, note that exchanges could be done fairly quickly and cheaply and that this was a facilitator in their decision to get involved. It was also felt that it was less likely that people would hear about the scheme if they were not internet users and may not be able to register for the scheme even if they were. A few participants had registered for the exchange on behalf of family members who were not digitally literate.

**Table 3:** Resources CMOs

| Overarching CMO statement | Illustrative participant quotation |
| --- | --- |
| 1. If people have limited time or financial resources then they may be less likely to participate because they have little spare time or money to put into buying cards/materials for creating cards. | *"You know when you're sending it, you have to post and then you have to pay for the postage yourself? And I know that postage isn't a lot, but it depends on where you are in life and what's going on." P8* |
| 2. If people do not have internet access or are not confident internet users, then they may not be able to participate independently because they are not able to register for the scheme. | *"Obviously for people who are not technologically literate you know who may only have a smart phone you know and are not on a contract and just you know do a pay by use type of sim card, it might be more difficult to access to sign up." P2* |

#### 4. Culture

Participants noted that KbP may be more accessible or have more of a draw to certain groups or types of people (table 4). Some felt that the advertising for the project and the act of creating and sending a card with a message of good will felt more typically feminine and so likely encouraged women in particular to participate. Participants had mixed feelings about attached exchanges to cultural events such as Christmas; many believed that it may alienate people who did not celebrate that cultural event and felt that they themselves did not feel any particular benefit from having it attached to the event.

**Table 4:**
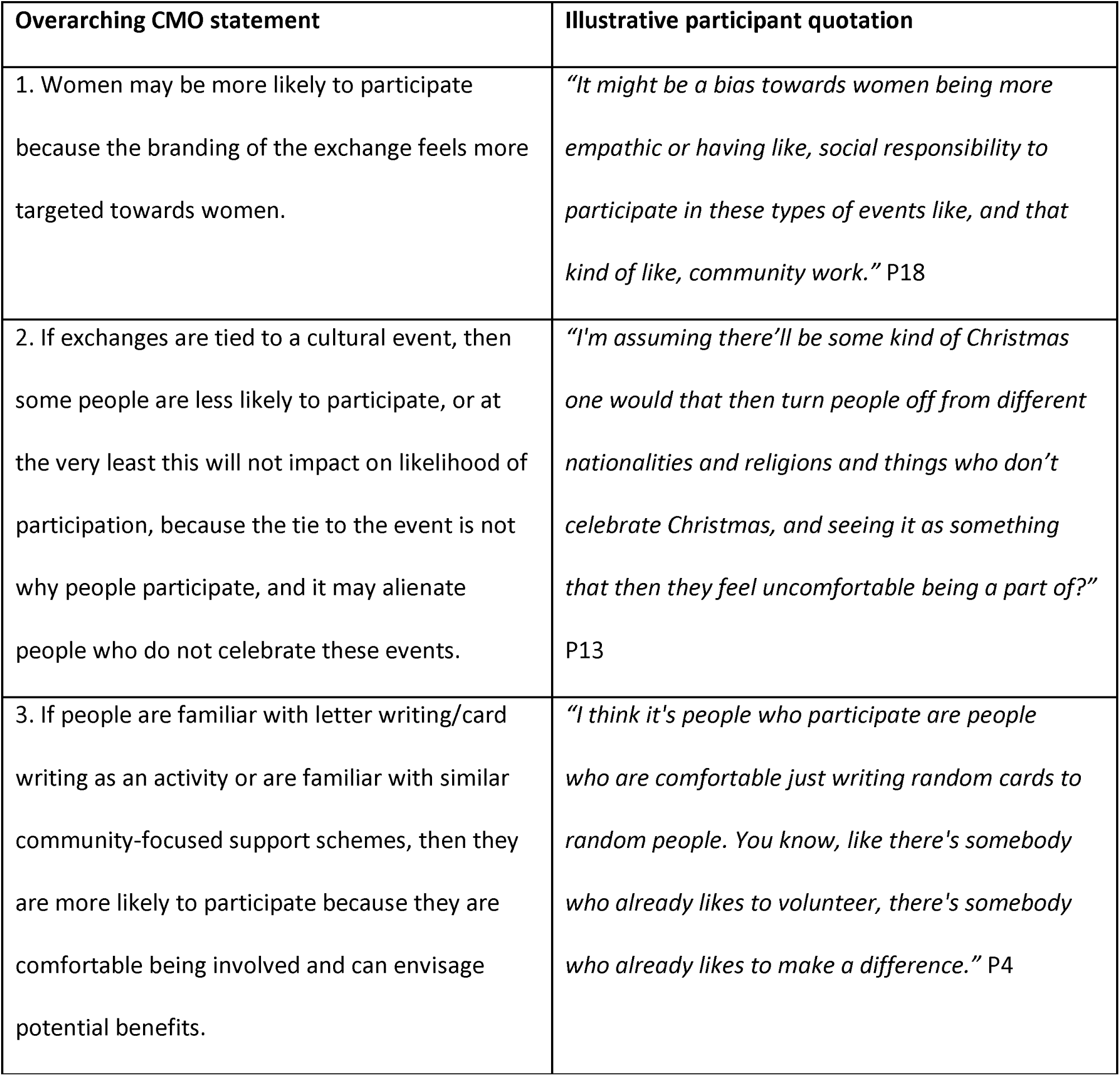
Culture CMOs

However, a smaller number of participants noted that it could be helpful given that people may feel lonelier at those times of the year and therefore may get more benefit from receiving post.

Participants with experience of card/letter-writing or community-type programs may be more drawn to the exchange because it was something familiar to them, and they signed up as they could envisage the benefit that it could bring. Alternatively, it was suggested that people who did not have much trust in their community, or who were not confident with English or used to card/letter sending could be less likely to participate.

#### 5. Giving post

Many participants were inspired by others to take part in exchanges, particularly through social media.

As well as being an effective mechanism for recruiting participants, it could also serve as a guide or a source of inspiration for what to write or create inside their card (table 5). This was particularly true for those taking part in their first exchange or those who found it difficult to know what to make or write.

**Table 5:** Giving post CMOs

| Overarching CMO statement | Illustrative participant quotation |
| --- | --- |
| 1. If people see images of diverse types of mail posted online (e.g., social media) or speak to others about what to send, then they may find it easier to create mail, because others' creations and experiences provide inspiration and ideas for their own creations. | *"I looked on the hashtag [#KindnessByPost] of it to see what sorts of things other people had written, to like get a bit of inspiration from that which helped." [P15]* |
| 2. If people do not know anything about the person they are sending mail to, then creating mail may be particularly effective in boosting mood, because creating mail becomes an introspective and reflective exercise, in which people imagine what their recipient would like to receive based on their own experiences. | *"You know I wasn't like giving an update on my life that I would for an aunt or something like that or trying to think about what's this specific person that I know well going to find interesting when you know finding things to send them or stuff like that. It is very much about a bit of a yeah, a leap of faith kind of thing and just, I guess the opportunity to think* |
|  | *about a hypothetical other person who you don't know anything about is quite nice and to do that in a positive way."* [P7] |
| 3. If people create their mail themselves, then this is an enjoyable experience with an opportunity for self-expression and learning, because it is a reflective and emotional process where one engages creatively with the process. | *"I've felt really good benefits from it because I carved out that time to actually craft something or draw something so that was my bit of me time away from everything else as well, to take time out to actually make that thing to send to somebody, so it gave me that permission to use that time in that way as well."* [P6] |

Anonymity was commonly discussed by participants. The majority of those who talked about anonymity felt that it enhanced their experiences of giving mail, and improved their mood because participants imagined what their recipient would like to receive based on their own experiences. This was experienced as an introspective and reflective exercise, whereby the anonymity of the recipient enabled freedom in what they created and why, and participants often found themselves writing things that they themselves wanted to hear. However, a smaller number of participants felt that they struggled with not knowing who would receive their card as they found it hard to determine what would be appropriate to send and what would have the greatest benefit for them.

Participants described different dimensions to their creative experiences when making cards. For some, it prompted moments of enjoyment and allowed for self-expression. For others, processes of creativity involved learning new creative techniques which were sometimes challenging, resulting in feelings of accomplishment. On the other hand, there were some participants who felt that the creative process of making a card was too challenging, and some who felt perfectionist tendencies such that ‘getting it right’ led to worry. Their preference was for buying a card and focusing on what to write inside it.

#### 6. Receiving post

Words of support and encouragement are a common feature in the cards. Participants found that receiving mail could boost positive well-being, feeling that receiving a personal card or letter in the exchange was something that stood out from their usual routine and usual received post (table 6). It was also tangible proof that somebody cared enough to send them post, raising their sense of self-worth. A number of participants felt that it was something that could also offer a protective effect against negative mood, as the memory of the exchange served as a reminder of the kindness of others if they experienced or saw something negative: a reminder that there is good in the world when times seemed dark.

**Table 6:** receiving post CMOs

| CMO | Quotation |
| --- | --- |
| If post is received in the exchange, then it can protect or boost positive wellbeing, because it is enjoyable to receive post, and it is proof of the kindness of others. | *"It was exciting to get something in the post because you don't get much apart from bills or horrible things that you don't want or junk mail, so it was just really, really nice."* P3 |
| If people do not receive post in the exchange, then their wellbeing may be negatively impacted, because they feel left out and do not feel comfortable enough to request a back-up card. | *"I did get missed out on one project and I was surprised by how hurt I felt so that's the only downside I've had."* P2 |
| If people do not receive post in the exchange, then their wellbeing may not be negatively impacted, because their benefit comes primarily from sending post and being part of the community, and they do not feel as though they | *"I never got one back and that was okay, you know, I filled out the form and I was like, well, that's not why I'm doing it. I'm doing it to make somebody else feel good and that is what is important."* P4 |
| need their own card or letter. |  |

Several participants spoke about not receiving post in an exchange in the past. Some felt that this had negatively impacted them, making them feel unimportant. Other participants felt that their primary motivation for taking part was the chance to share kindness with or help somebody else, and so they did not see receiving post as an important part of their experience. All participants had the option of using a back-up system, where people can report not receiving a card through the website and be sent one by a volunteer. Some people did not feel comfortable making use of this system, in some cases despite being negatively affected by not receiving post, as they did not want to be a burden on the system.

#### 7. Content of received post

Beyond the broader impact of receiving post in the exchange, the content and type of post received influenced participants (table 7). Participants often recognised the effort that was put into letter-writing or crafting a card, and the knowledge that this effort had been made for them added to the joy of receiving post. Meanwhile, a low effort card, such as a shop bought one without a handwritten message, could be disappointing or otherwise produce a negative response from the receiver. Comparisons were often drawn between the ease of sending a text or an email with warm wishes as opposed to the time invested in writing a letter or making a card and then posting it. Similarly, post that felt personal, either by the sender writing about their life, or by engaging with where the recipient lived (the only known thing about the recipient), added to a sense of connection and effort on the part of the sender.

**Table 7:** Content of received post CMOs

| Overarching CMO statement | Illustrative participant quotation |
| --- | --- |
| If people receive post, that has clearly had effort put into it, then this can feel particularly | *“if you send something physical like a letter it shows that someone’s taken that effort to do that, it’s not* |
| meaningful, because they appreciate the time and effort that the sender put into doing something kind for a stranger. | *just like Ah I can just send a text, or I can just talk to somebody through a screen. You've actually taken the time to go out, get a letter or a card or whatever, put a bit of thought into what you're writing, and purposely sent it to someone."* P5 |
| If you do not know anything about the person you are exchanging with, then you can still get a lot of value from receiving your card, because just being kind to people together is enough of a connection. | *"You don't know any information apart from a first name and a postal address for somebody, it means that it's not about all the things we've got in common, it's just about being a human being and by virtue of being another human being, and being able to say something that matters to somebody."* P1 |
| If what is written in received mail is applicable to your current circumstances, then this can feel particularly meaningful/helpful to wellbeing, because it can feel like it was the right message at the right time; a deeper connection with the mail and the person who wrote it. | *"There is something uncanny and like surprisingly powerful I think, especially if you're – if you're in a receptive mood and there's a lot of synchronicity that people report, so they'll report "Oh my goodness, the message just came at exactly the right time" and like "It was amazing how it spoke to certain things, it was just what I needed to hear," people say things like that."* P1 |
| If the sender has made references to particular faiths, beliefs, or to the personality, feelings, or experiences of the recipient, then this can detract from the joy of receiving mail, because the recipient can feel alienated if they do not | *"[there] was a religious inference, or a religious reference, in there ... and that just made me a feel a bit uneasy... it did have an alienating effect on me, and it could have an alienating effect on other people as well who are outside of the kind of white* |
| ring true for them or they are viewed as problematic. | *Christian heteronormative majority.” P12* |

Some participants received messages in their post that felt highly applicable to their current personal circumstances; as the sender could not have known about this, these messages could feel particularly meaningful and gave a sense that there was a special connection between them and the sender. Participants did, however, often feel that just the act of sending and receiving post connected them with others in the exchange, as they could identify with the common desire to share kindness with others. A lack of personal information did not detract from this.

A small number of participants had a negative experience of senders making references to specific beliefs, such as religious beliefs, or to the personality, experiences, or feelings of the recipient; as the anonymity of the exchange meant that the sender could not know that the recipient would identify with/value those words/thoughts and assumptions may have been made. For some, this detracted from the positive impact being involved in the exchange may have had.

#### 8. Community

Many participants spoke about the sense of community that involvement in exchanges brought about (table 8). Social media was a particularly powerful catalyst for this, as seeing people post about KbP exchanges reinforced the knowledge that there were many people participating in the exchange, and not just those who sent or received their cards.

**Table 8:** Community CMOs

| Overarching CMO statement | Illustrative participant quotation |
| --- | --- |
| 1. If someone shares a card they have made or received on social media, then it may reinforce their sense of community and belongingness and enhance wellbeing, because it reinforces being part of a wider, collective activity and counteracts more usual negative experiences of social media. | *"You see on social media there's like, you can see this you know 10,15, 100 other people exchanging and that adds to the warm fuzzies."* [P19] |
| 2. If people post the card they received or see the card they made on social media, then it may add to wellbeing, because it lets the other person know the card was received and valued. | *"Because I thought the card, the person has made a lot of effort into that card so it would be nice if they could see that it has been on Instagram."* [P9] |
| 3. If people involve others in the card exchange (e.g. their families or colleagues) then it can add to wellbeing and a sense of connectedness, because it involves doing a fun activity together and pleasure from seeing others' enjoyment in the process. | *"That was fun to have that little community at work participate."* [P1] |
| 4. If people share some information in their card about their culture or the area where they live, | *"It could be beneficial for people who live in more homogenous communities to get post letters,* |
| then this may increase people's sense of wellbeing, because they feel connected and part of a community with people they might not otherwise have contact with. | *cards from people from you know different communities, I think that's really beneficial, you know when I see a non-Anglo Saxon name I'm like oh cool, like you know I'm writing to someone who I know is from a different ethnic background, so yes I think it could be helpful on that front, yes."*<br>[P2] |

Participants who shared their post online also often made additional connections with others who interacted with their posts, finding that it was a way of sharing the act of kindness with their friends and followers. Participants were aware that shared images of post may connect senders and recipients to each other; many felt that this was positive as it demonstrated that post was appreciated and valued. Some would search social media platforms to see if their post had been shared. Seeing post could make the exchange feel more “real” as it made senders and recipients less anonymous. However, social media was not universally valued; some participants did not use social media at all, and one participant said that detracted slightly from the exchange feeling personal and special and other participants refrained from sharing received post online as they were conscious that the sender had not consented to their post being shared with a wide audience.

A few participants derived a sense of connection by creating post or signing up to exchanges with friends, family, or colleagues. Others valued receiving post where they learnt something about the sender and their community, as they were introduced to a new place or culture in the UK that they may not have known about. However, this is contrasted with the insight (from theme 7) that references to specific beliefs/faiths could result in a negative experience for the recipient. Together they suggest that people enjoy learning about others in the exchange but dislike the feeling that the sender has made presumptions about their beliefs, values, or personality.

#### 9. Long-term personal impact

Participants talked about the impact of participation beyond the exchange period (table 9). Post was often kept; some participants displayed cards prominently, whilst others kept them stored but would access them when they wanted to improve their mood. Post served as a reminder of the kindness of others, and messages in the post continued to benefit participants. For some participants who took part in more than one exchange, they reflected that in the time between exchanges, they were more likely to look for and remember positive messages that they could share in their next exchange, making them more likely to internalise these messages. Being part of multiple exchanges also seemed to increase the sense of community provided by KbP, and participants benefited from having future exchanges to look forward to; both the sharing of kindness and the opportunity for creative and reflective activity. A few participants also felt that involvement in exchanges further prompted them to engage in similar creative or kind acts.

**Table 9:** Long-term impact CMOs

| Overarching CMO statement | Illustrative participant quotation |
| --- | --- |
| 1. If received mail is kept and displayed prominently in the home, then it may sustain wellbeing, because it acts as a day-to-day reminder of the kindness of others; useful affirmations or kindness tips, and the KbP community. | *"So, I actually look at it every day now...and now this is like a positive impact that kind of carries on, because I always have it next to me so I can always look at it every time I'm feeling like down." [P8]* |
| 2. If someone gets involved in future exchanges or plans to, then this may sustain wellbeing, because the person attends more to examples of kindness or positive messages which could go in future cards, and a mindset of positivity and kindness is developed. | *"In between card exchanges I'm always aware of new ideas that I would be able to use for the next exchange, so if I read something or someone says something or I see something or hear something and I say oh I could probably use that, I'll put a little note in my phone for next card exchange, like a quote or something that I think that made me feel happy that I hope to pass on to whoever the recipient would be." [P1]* |
| 3. If KbP exchanges are frequent and someone | *"[Taking part in KbP exchanges] are really nice* |
| takes part regularly, then additional benefits to wellbeing may accrue, because they start to feel part of a community, and taking part in the card exchanges becomes increasingly valued and important. | *patterns, nice ways to be thinking I think, and I really appreciate that opportunity to be in that particular headspace and I think more – that’s really useful more and more yes.” [P7]* |
| 4. If taking part in KbP prompts someone to continue with a creative activity like crafting or letter writing, then there may be additional benefits to wellbeing, because it builds confidence and leads to continued engagement with a rewarding creative activity. | *“This is actually a really good thing that came from the Kindness by Post was that it inspired me to write cards to random friends and family and tell them who they are to me and tell them that I love them.” [P4]* |

### Theory of Change model

Figure 1 presents our finalised Theory of Change model, reflecting our programme theory of KbP. It details the participant-level pathways that lead to specific outcomes. The model is based on the overarching CMOs presented in previous sections, with statements and causal links summarised for simplicity. During the analysis process, the number of outcomes increased as the understanding of the nuanced impact of the intervention became clearer. The number of barriers increased as unintended consequences became more apparent, and the number of mechanisms increased and became more specific to the distinct elements of giving post and receiving post.

**Figure 1:**
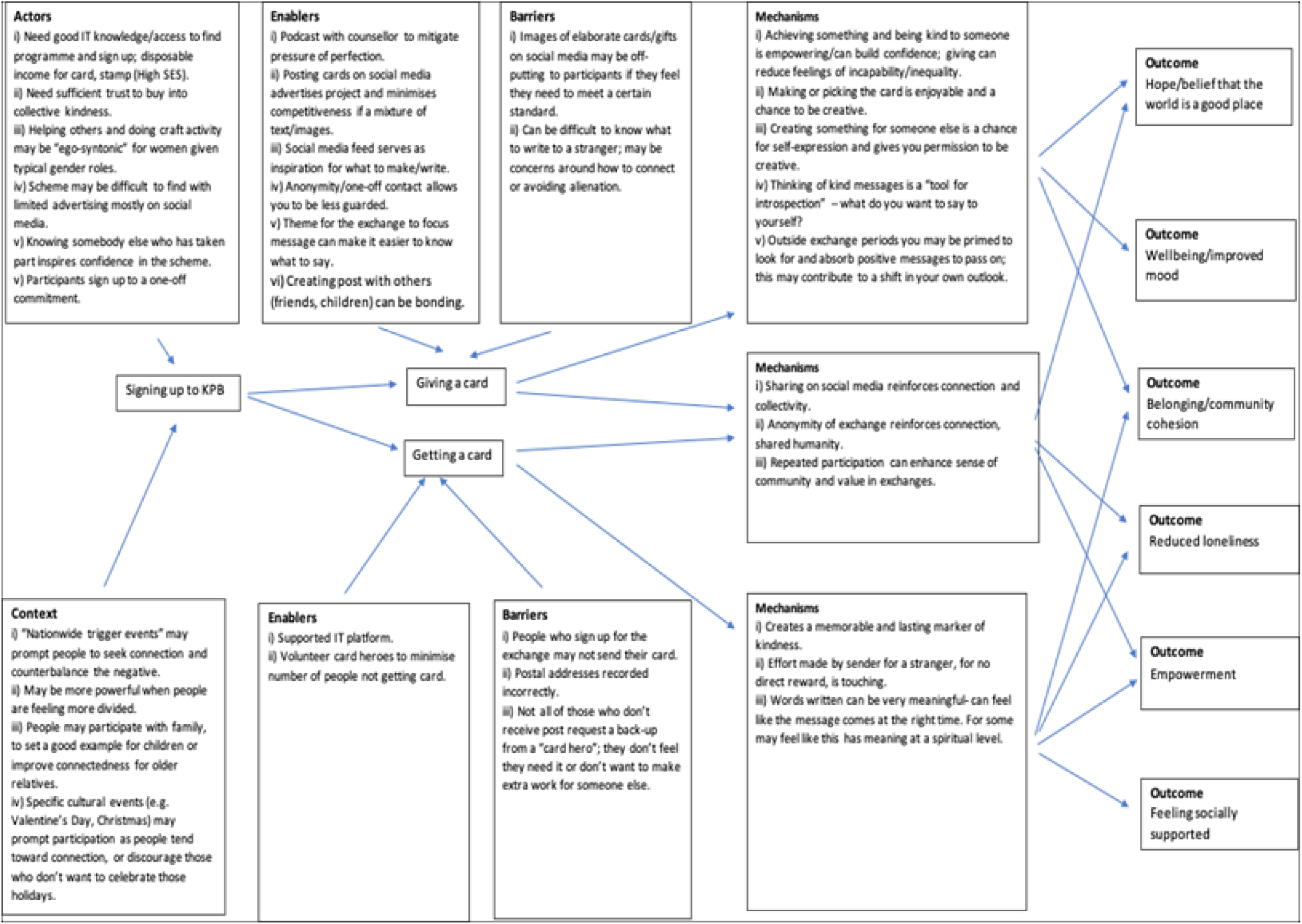
Theory of Change model for KbP intervention

## Discussion

### Summary

In this study, we developed a programme theory for a participatory public health intervention Acts of Kindness exchange “#KindnessByPost”. KbP produced a range of outcomes for participants, both during and after the exchange period. Our Theory of Change model presented above offers a clear programme theory that explains the contexts and mechanisms that lead to specific outcomes. Both giving and receiving post, as well as the sense of belonging to a community offered outcomes related to positive mood and connectedness. Connection with a stranger as opposed to exchanging messages of kindness with a friend had a particularly salient impact on participants, as did the physical nature of sent and received post. Causal pathways suggest that there are a range of contexts leading through to positive outcomes in which people are willing to participate, and so the intervention is likely applicable across communities and settings.

At a collective level, wider culture (particularly specific events and familiarity with post as a form of expression) influenced participation and at the individual level. Involvement typically required some level of digital literacy to find the intervention and register for it. KbP was also perceived by some also more geared towards women through the presentation of the website; and sending cards by post is generally a more typical activity for women than men in a UK context where women buy 80% of the greeting cards sold in the UK [18].

### Findings in context of existing research

Our findings and consequent programme theory are also consistent with an earlier evaluation of KbP [12], which suggested that potential mechanisms for the intervention were pleasure in making and sending cards, individual fulfilment and appreciating other’s thoughts and behaviours. Furthermore, quantitative findings from both that study as well as a more recent and larger evaluation [13] indicate that participation in KbP improved wellbeing and loneliness, and, in Wang et al. [12], increased feelings of belonging. Our findings reflect this: most participants in our study reported positive experiences of KbP, including that they enjoyed receiving something tangible in the post and it reminded them of kindness of strangers, they appreciated the sender making an effort with their card, that they liked feeling part of a community and looking at other’s posts on social media, and that they often enjoyed the creative process of making a card themselves. These improved participants’ feelings of wellbeing and connectedness with others. We also heard reports of converse experiences from a few participants: that not receiving a card, receiving a low effort card or one with a message that they found alienating, or finding it stressful to create a card all could have a negative impact.

Also reflecting our finding that most participants tended to value sending post over receiving post, Wang et al. (2022) found no significant difference in improvement to wellbeing between those who only sent post and those who sent and received post. These results suggest the offering of kindness was an important mechanism in positive outcomes and support the idea that the act of helping others without the expectation of a reciprocal act is known to benefit wellbeing [19, 20].

The results of Le Novere et al. [13] also indicate that these positive benefits are sustained at least 3 months later, which again are reflected and explored in our results. Participants spoke about how they sustain the benefits of KbP between exchanges by keep the post they received, displaying it prominently or storing it somewhere easily accessible; by planning for future exchanges and looking out for inspiration for the cards they will create; and by regularly taking part in exchanges such that benefits accrue over time and the experience of participating becomes more valued over time.

Each of the principal elements of the KbP intervention: sharing kindness, making/writing the card, receiving kindness, and being part of a community were valued by our participants and elicited perceived positive outcomes are broadly consistent with evidence regarding the association between community engagement and engagement with the arts with a positive effect on wellbeing [6, 8]. Such evidence has been pulled together by Fancourt et al. [7] in the Multi-level Mechanisms Framework, which is a framework that describes the associations between leisure activities and health/wellbeing. In this study, there were clear psychological processes leading to positive outcomes that are included in this framework including the building of an identity with the KbP community, the positive affective experience of receiving post, the creativity of making/writing the cards, and post kept as mementos to improve mood beyond the exchange period.

### Strengths and limitations

Using a realist approach to data collection and analysis allowed us to generate a nuanced understanding of the potential causal relationships between the context, mechanisms, and outcomes of the intervention, acknowledging that an intervention cannot be expected to produce the same outcomes for every participant in every setting. Producing initial programme theories based on the KbP executive’s intended outcomes for the intervention allowed us to test and better understand their proposed causal mechanisms from the perspectives of intervention recipients. As programme theories were developed collaboratively within the working group, and refined and consolidated in partnership with participants, results are highly corroborated.

The sample was relatively diverse in that it included participants of different ethnicities, ages, and sex, as well as those who had taken part in just one exchange and those who had taken part in many. However, the demographics of those that take part in KbP are strongly White British and female [13]. Similarly, the sample in this study was predominantly White British and female. We believe it would be useful to explore the perspectives of male and ethnic minority participants further to understand better why this is, and how KbP and other Acts of Kindness interventions could be tailored or adapted to other populations.

As is typical for research of this nature, recruiting participants via official project channels was necessary to comply with data protection, but this may have put off exchange participants who had a less positive experience of the exchange. Similarly, it was not within the scope of the study to interview people who were aware of KbP but had not taken part in an exchange. As such, our sample may have offered us a more positive view of the intervention than may be expected from the wider population, and we may not have been able to capture important barriers to participation or more acceptable alternative approaches from the experiences of those who chose not to take part.

The digital promotion of KbP was at the forefront, spread between the website, social media, and email communications. Those who are digitally excluded may not be adequately represented yet they may also be more likely to be in the habit of sending cards and letters by post, and the Covid-19 pandemic restrictions on in-person meetings and the closure of community venues may also have limited our outreach to this population.

### Recommendations for research and practice

The study supports the potential value of participatory public health projects for improving wellbeing. KbP and similar interventions present highly scalable and low-resource ways to improve public mental wellbeing; given the promising and growing body of evidence, public health bodies should consider supporting widespread implementation and evaluation of interventions designed to improve connectedness and/or support engagement with arts to assess feasibility and impact on a large scale. Support could range from general promotion and publicity for such schemes, to funding for community groups and social hubs to support populations to initially engage (help to register, card making workshops, covering cost of postage etc.) with a focus on those most likely to benefit or those who are less likely to be able to participate independently.

For KbP, further quantitative evaluation could provide corroboration of the causal mechanisms and outcomes identified in this study. Additionally, future studies could establish whether the intervention could be particularly effective for participants with lower wellbeing or mental health difficulties, with samples in evaluations so far being drawn from the general population. Previous research does show that arts interventions can be effective in improving wellbeing for those with diagnosed mental health conditions [10]. Another interesting topic is the spontaneous creation of groups to collectively participate in KbP, including work colleagues, classrooms, and families. Given the evidence in this study regarding the possible value of these groups to amplify and sustain improvements to wellbeing, we believe it would be valuable to investigate this phenomenon further, including studying the differences between collaborative community participation and individual’s participation. This could assist with how best to expand the model to a wider population including groups which may need some support to participate.

The low participation rates of men and ethnic minority groups in KbP exchanges [12, 13] is notable. Despite best efforts to purposively sample from these populations, we had limited success. From our sample, we have not been able to ascertain whether the intervention brings out the intended outcomes for these group, nor fully explore the acceptability of the intervention for these groups. Further research with under-represented groups to explore the perceptions of KbP and barriers to participation in the intervention should take place; it may be that suitable adaptations to KbP or how it is publicised can be identified. During stakeholder discussions with community BAME health groups, it was suggested to one of the authors that distinct minority communities have their own ways of expressing kindness, such as gifting food or offers of help in the home. It may be that an inherent aspect of the KbP exchange design, such as the use of postal cards, mean that it initially has low appeal to some communities/groups who are culturally less likely to be card-senders. In this case, other Acts of Kindness exchange designs could be considered so as to appeal to distinct and diverse populations. Whilst it is unlikely that these measures could completely overcome structural health inequalities, we believe that the current low rates of engagement amongst these groups is important to consider, and opportunities to have more targeted designs or engagement activities be identified if feasible, especially where needed to avoid further marginalisation of already underserved groups.

## Conclusions

In general, participants in KbP experienced the intervention positively, reporting improved feelings of connectedness and wellbeing as intended by the KbP executive, reflective of quantitative evaluation of the project. Although some negative experiences were reported and included in our programme theory, it was notable these were generally uncommon, being reported by only a few participants. The Theory of Change model set out clear pathways through which the intervention created a range of positive outcomes for participants, including by creating positive affective experiences from receiving post, from creating post, and from the communities and groups that emerge around the KbP exchanges. KbP is an example of a highly scalable, simple, and cost-effective public health intervention that uses community and engagement with arts to improve wellbeing.

## Data Availability

The datasets used and/or analysed during the current study available from the corresponding author on reasonable request.

## Abbreviations

CMO: Context-mechanism-outcome statements
KbP: #KindnessByPost
UK: United Kingdom

## Declarations

### Ethics approval

This study was approved by the UCL Research Ethics Committee (ref: 14649/004)

### Consent to participate

People taking part in a #KindnessByPost exchange expressed interest in participating in the present study via an online form. They were contacted by a study team member who discussed the study with them and provided them with a participant information sheet outlining the aims of the study, that their participation was voluntary, that their data would be stored securely and anonymously, what would happen if they participate, and how to withdraw from the study or raise any concerns. Consent was taken at least 24 hours later prior to data collection interview.

### Consent for publication

Participants gave consent for their data to be published as part of a publication in a peer-reviewed journal during the outlined consent process outlined above.

### Authors’ contributions

LSR, BLE, HS, KF, and KW contributed to conceiving and designing the study, and LSR, BLE, KW, and HS planned the analytic approach. HS conducted the data collection and all authors were involved in conducting the analyses. HS led on writing the results and drafting the manuscript, with significant contributions from LSR. All authors contributed to drafting the initial manuscript, and all authors reviewed iterative complete drafts of the manuscript multiple times prior to submission.

## Funding

This project has been funded by the Loneliness & Social Isolation in Mental Health Research Network (549210/Rains), which is funded by UK Research and Innovation (UKRI) (Grant reference: ES/S004440/1) and their support is gratefully acknowledged. Any views expressed here are those of the project investigators and do not necessarily represent the views of the Loneliness & Social Isolation in Mental Health Research Network or UKRI.

## Acknowledgements

We would like to acknowledge the contribution made by Dr. Rebecca Jones, Marie Le Novere, and Dr. Caroline Clarke, who worked on the companion statistical and economic study and helped inform the present study at an early stage. We also would like to acknowledge the contribution of the Mental Health Collective, who are responsible for #KindnessbyPost and who helped with conceiving and recruiting to the present study.

## Appendix

Appendix 1: Final list of CMOs (Context-Mechanism-Outcome statements)

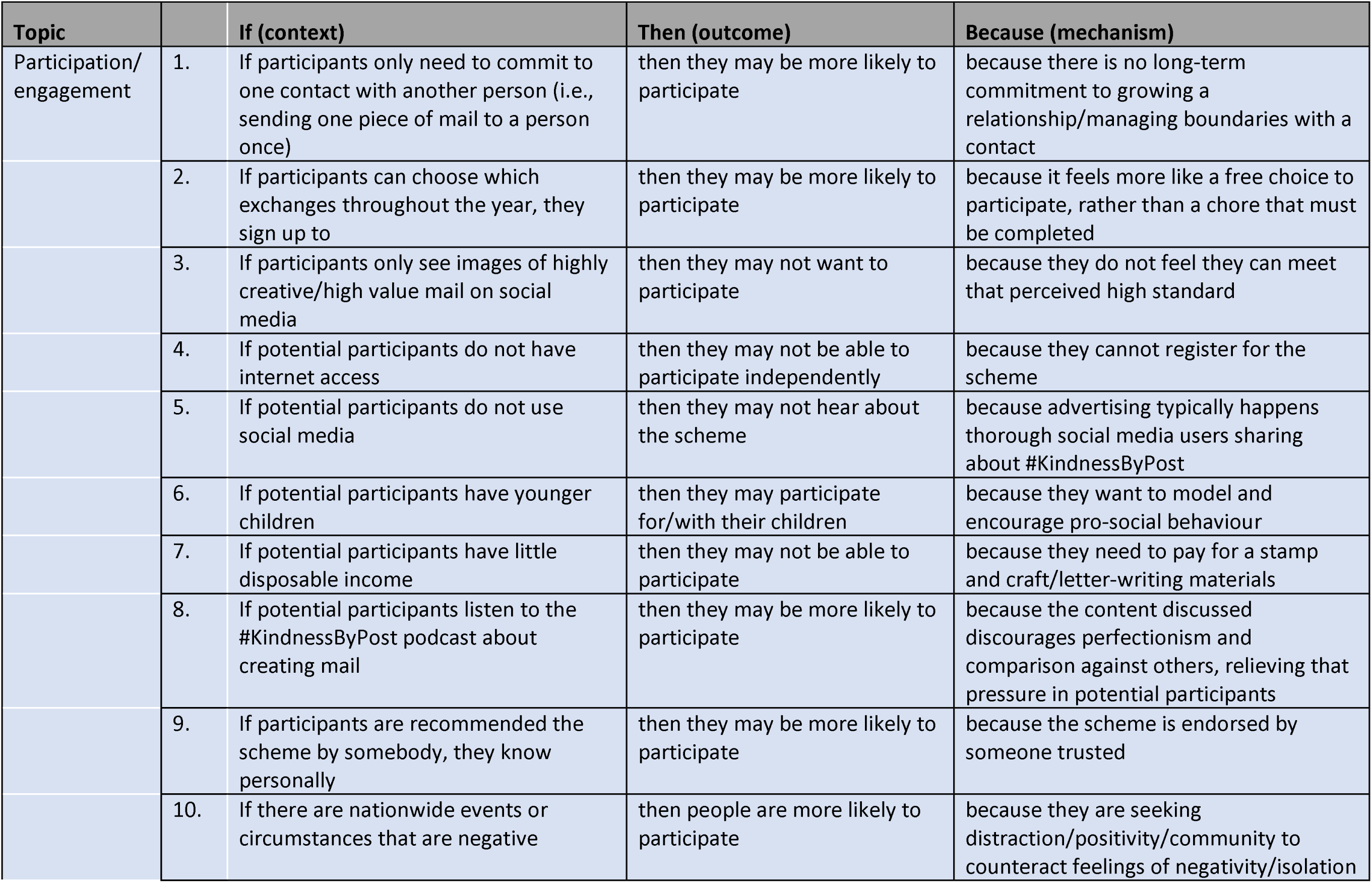
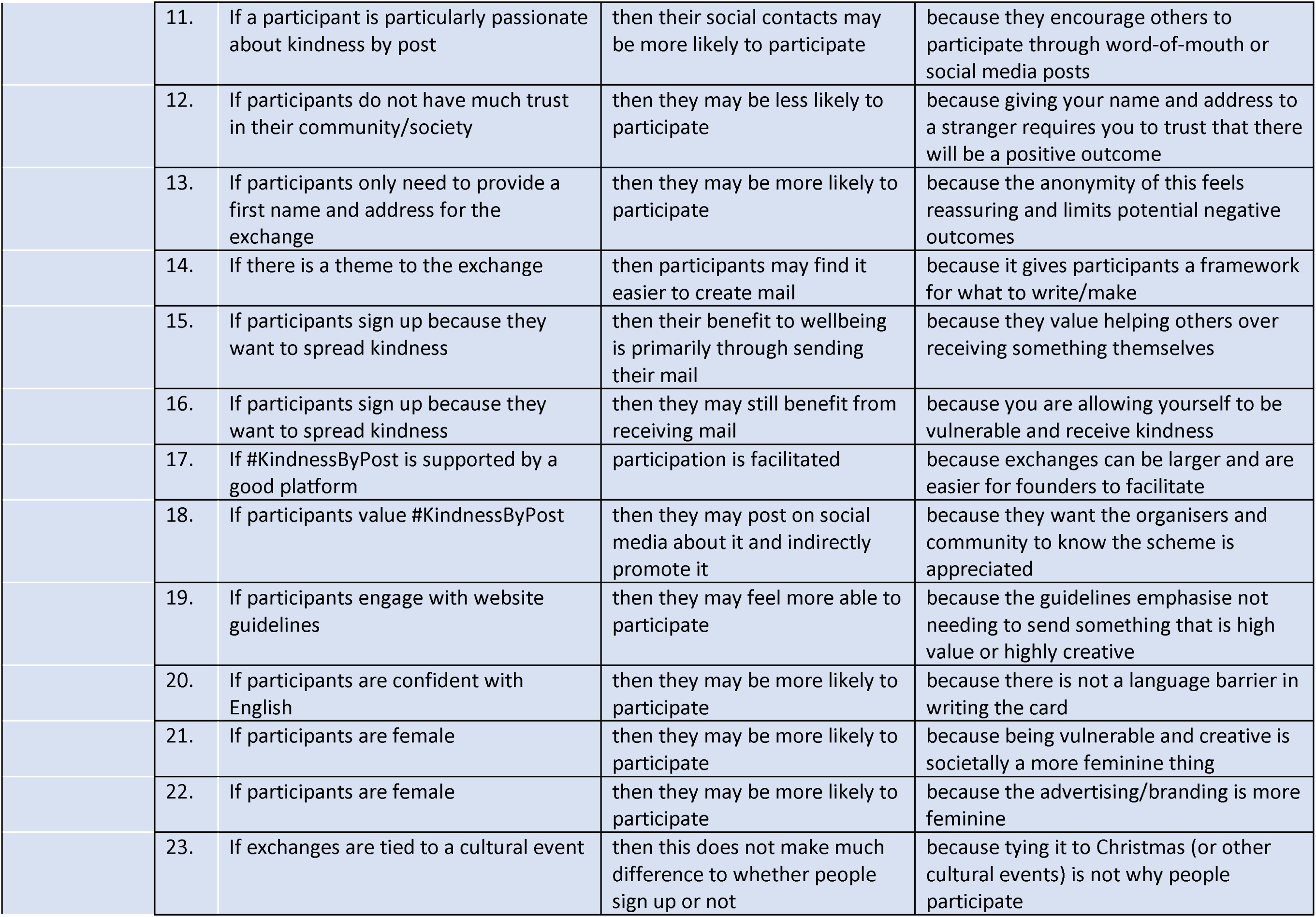
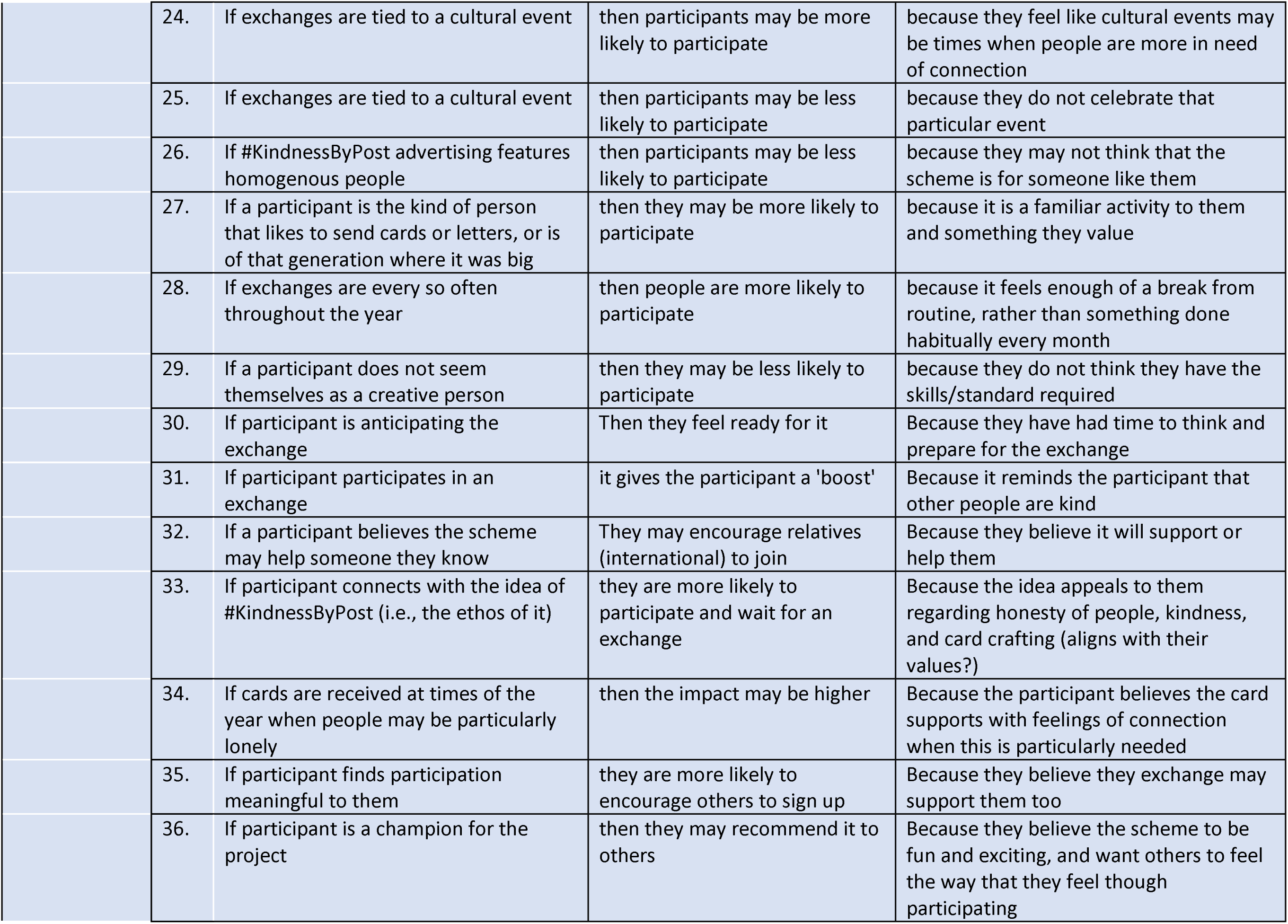
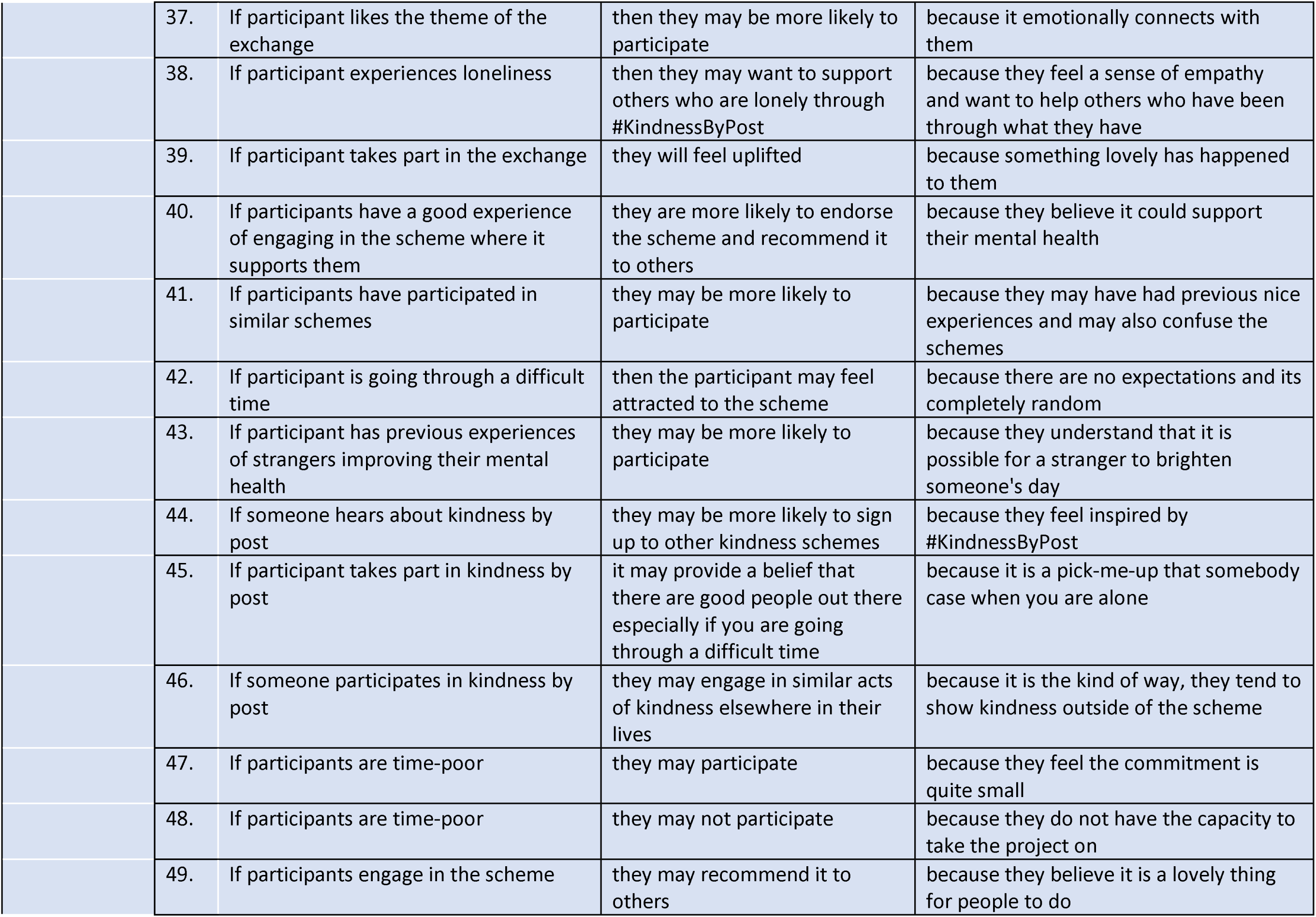
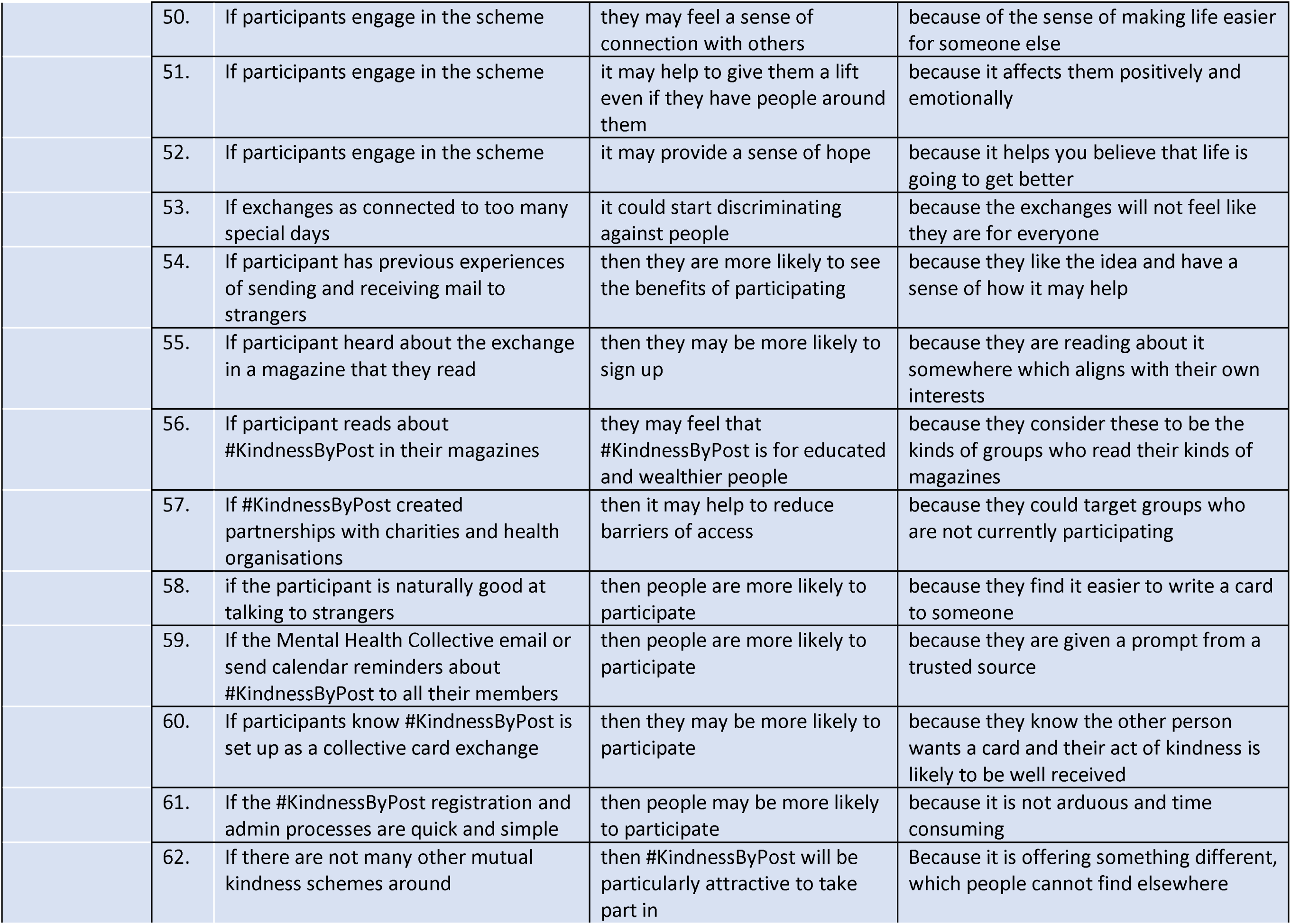
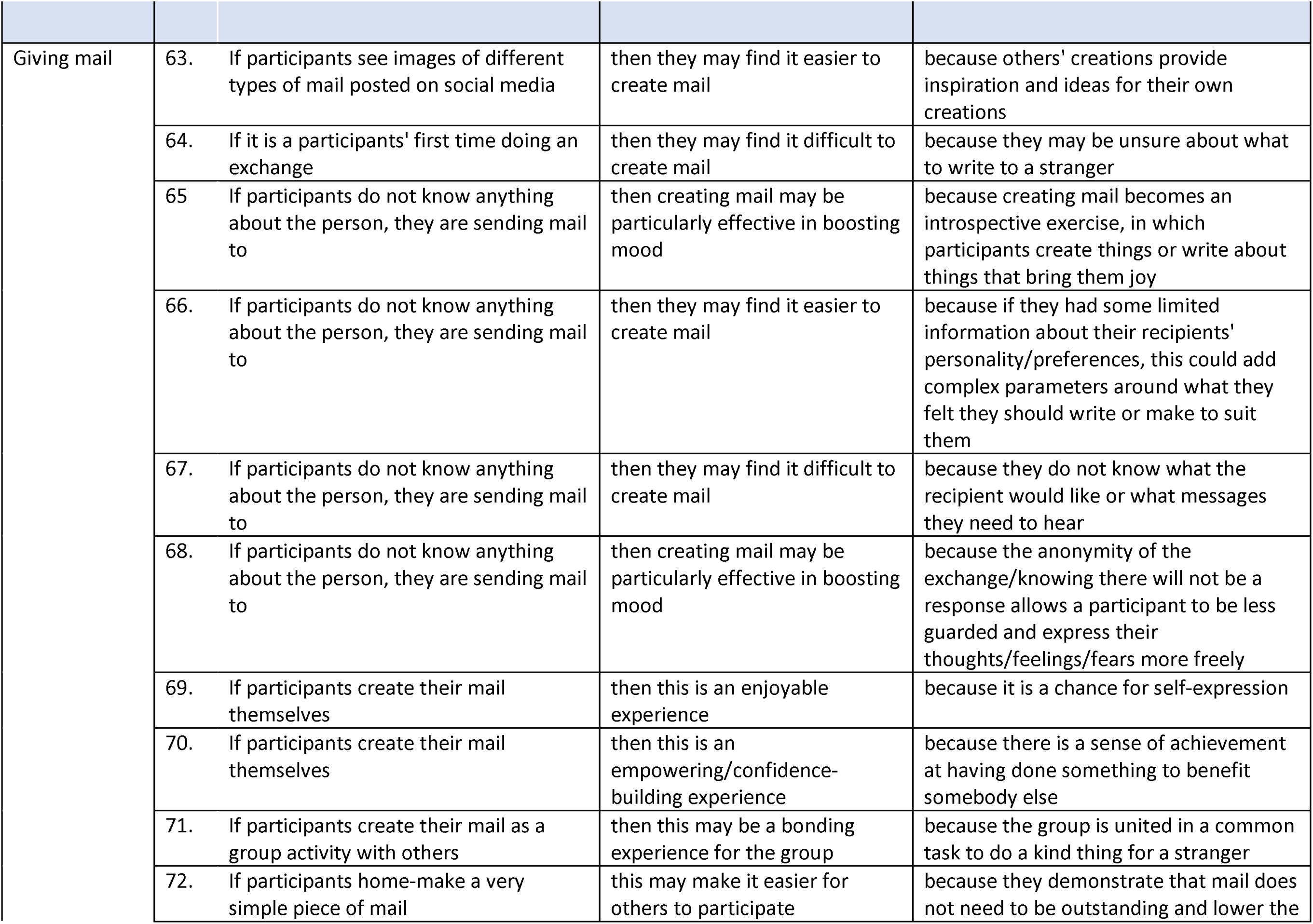
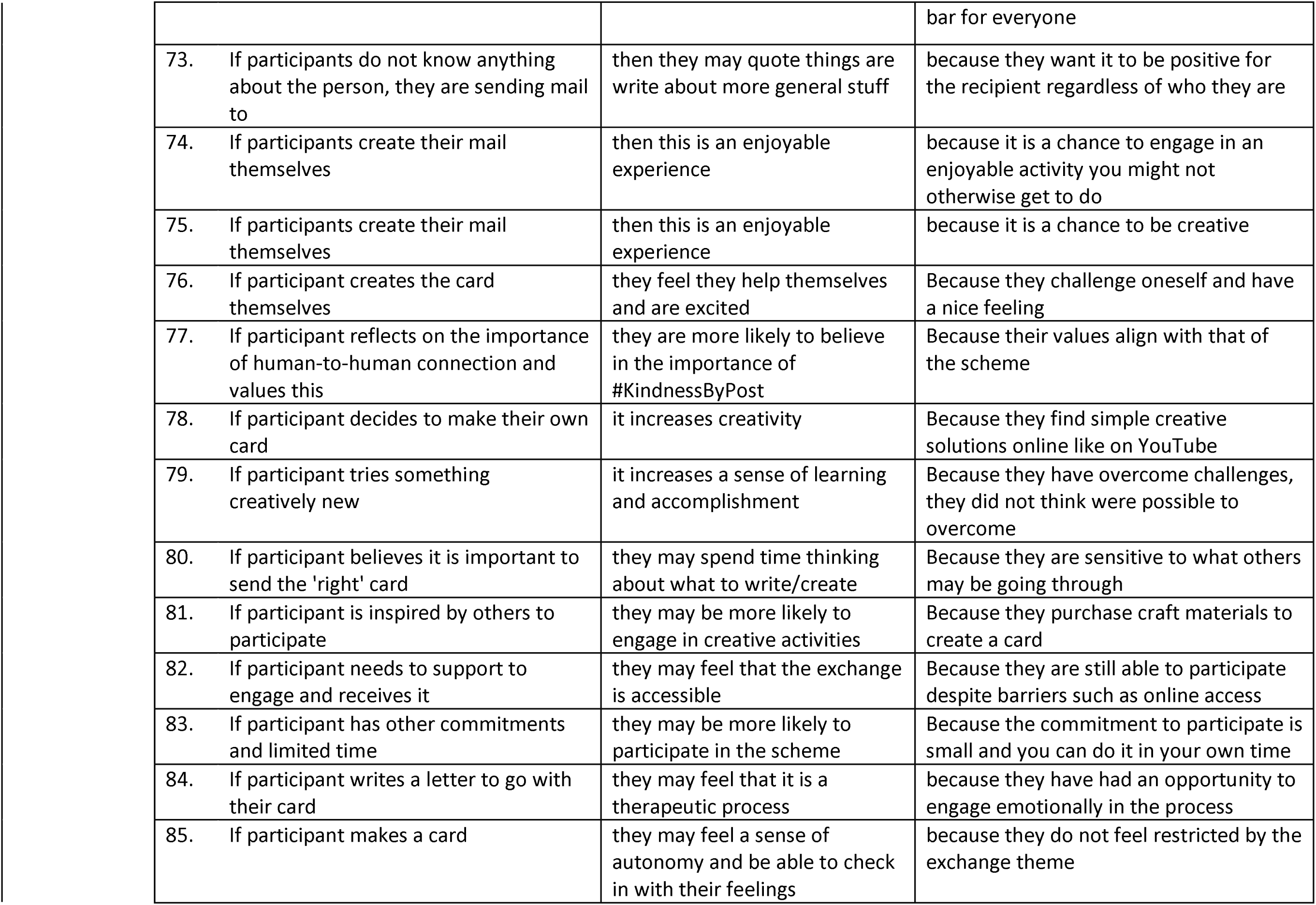
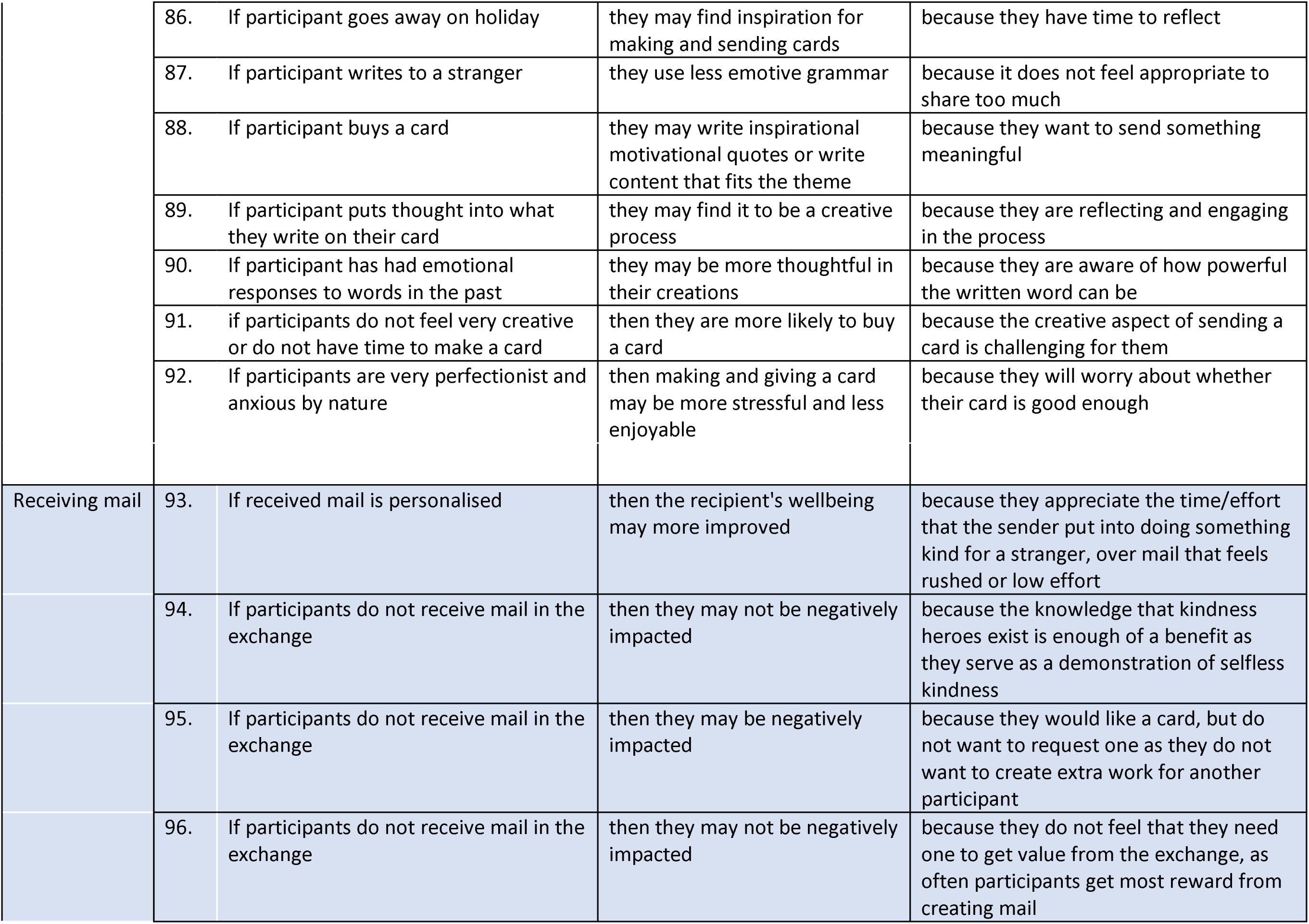
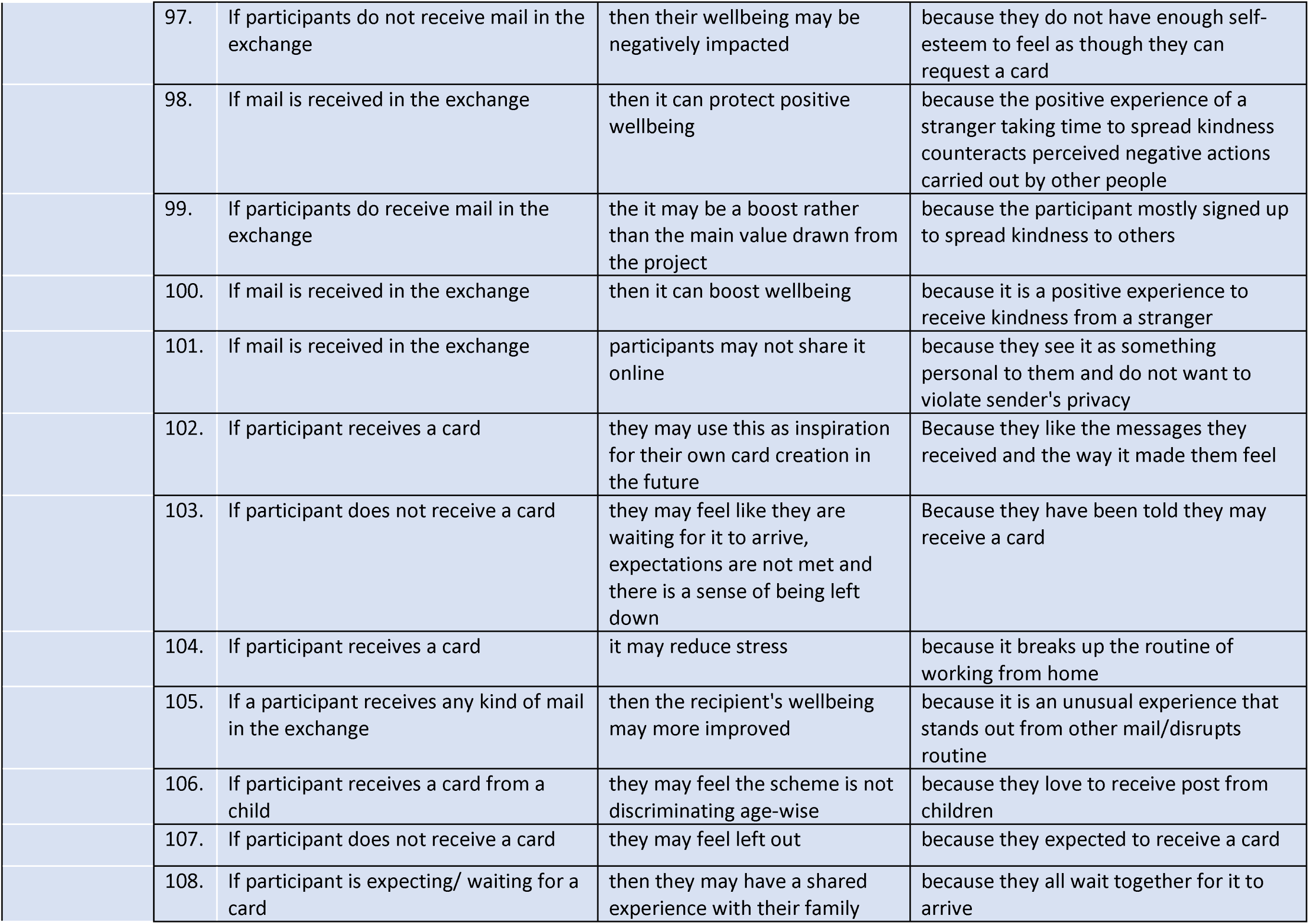
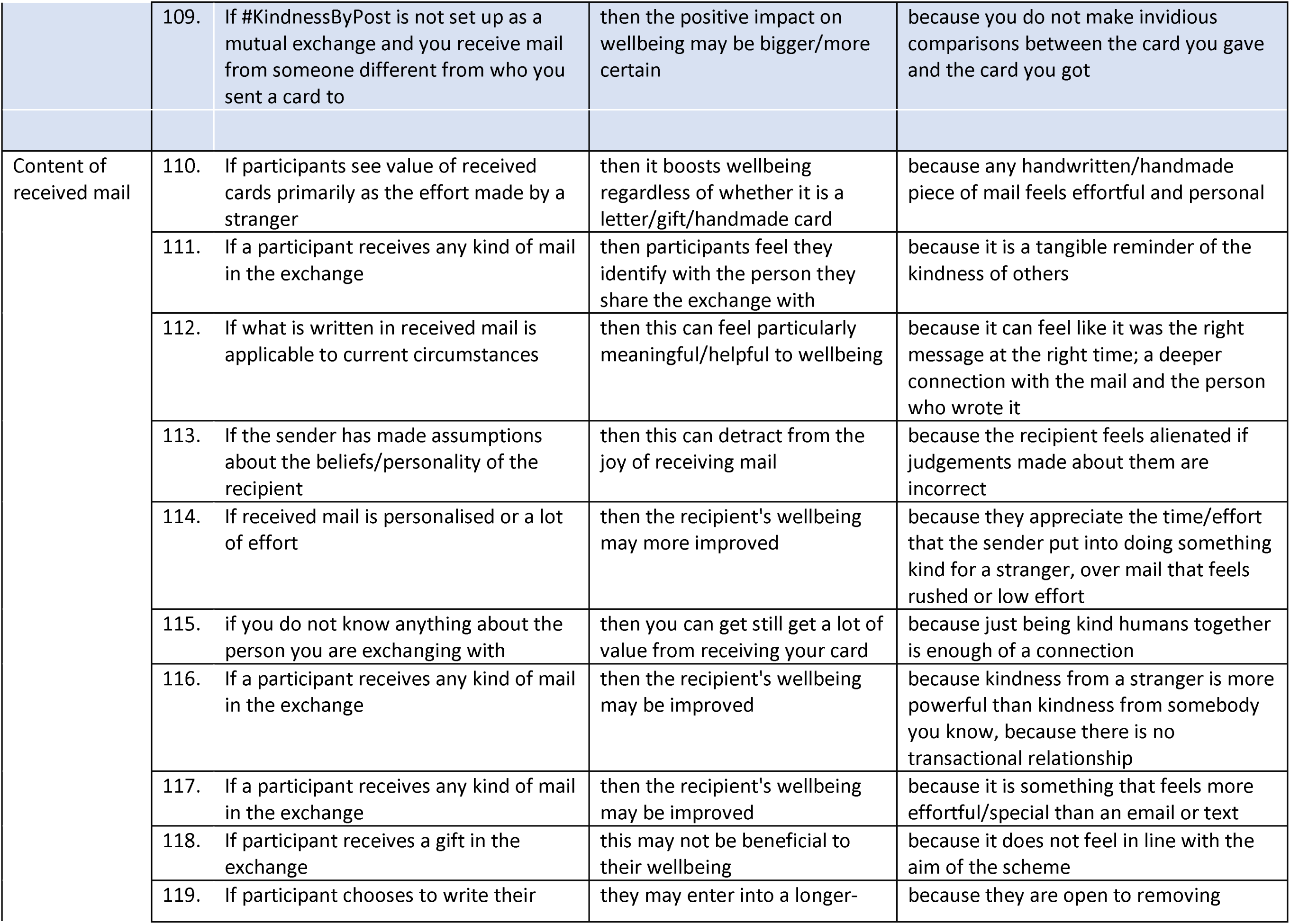
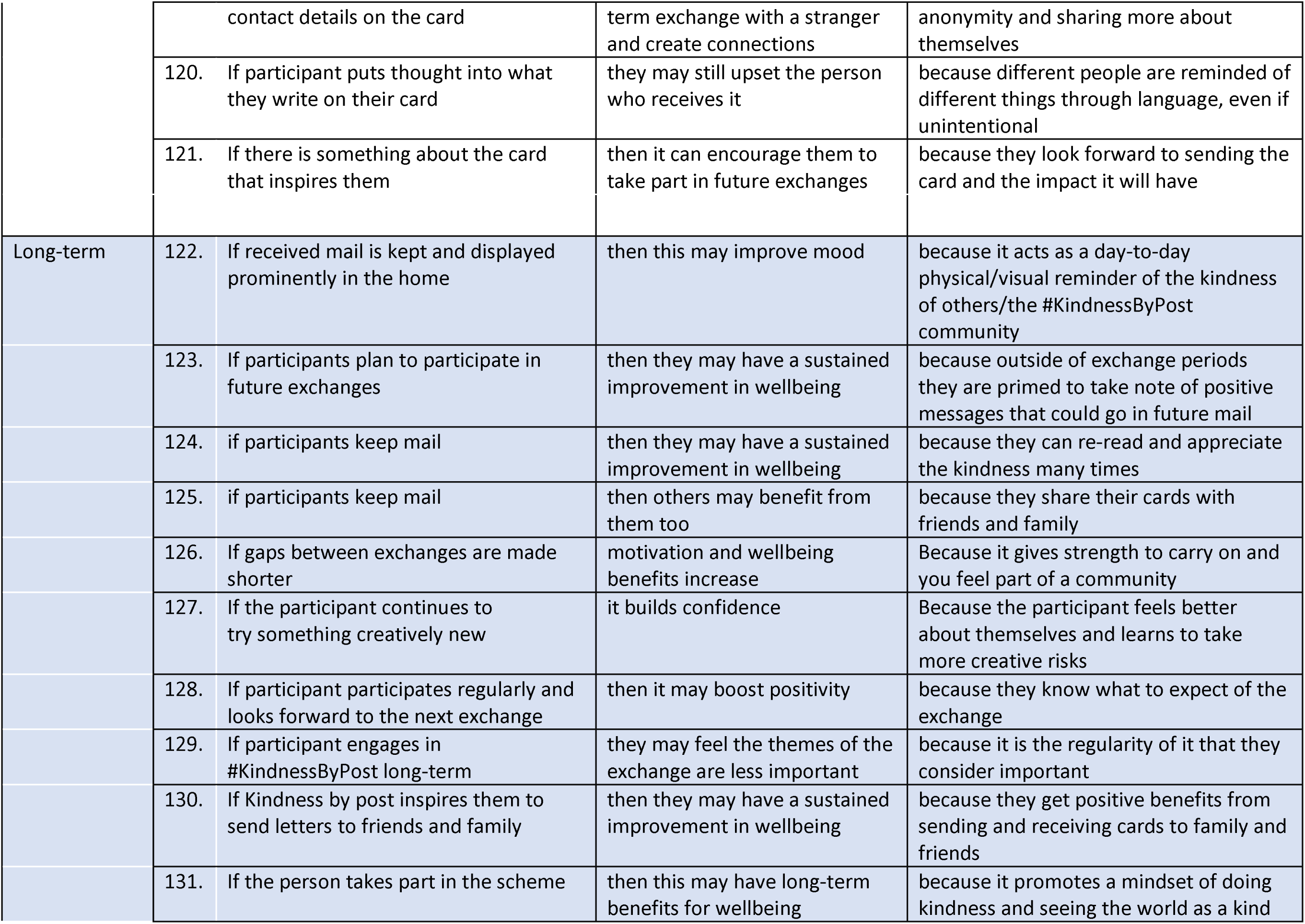
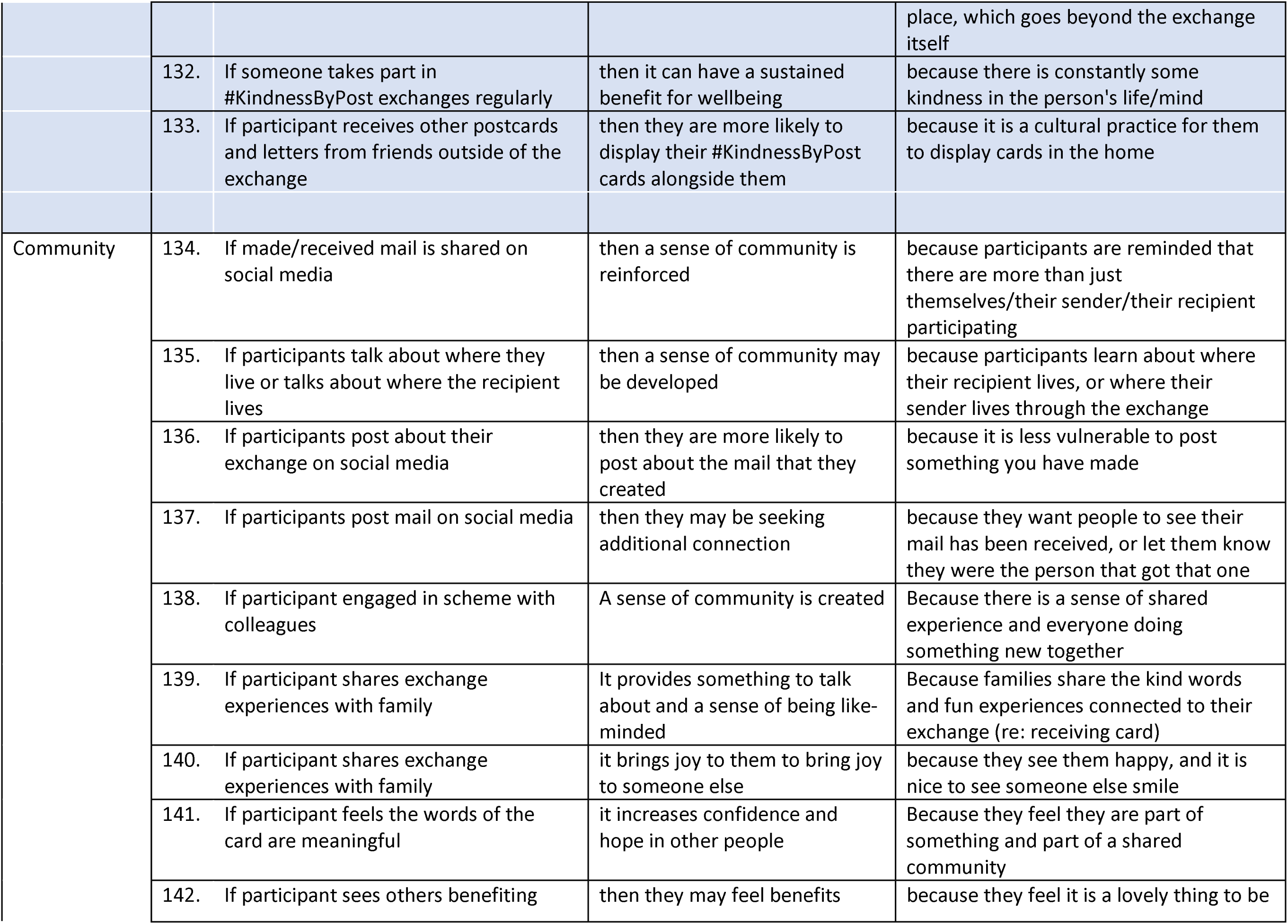
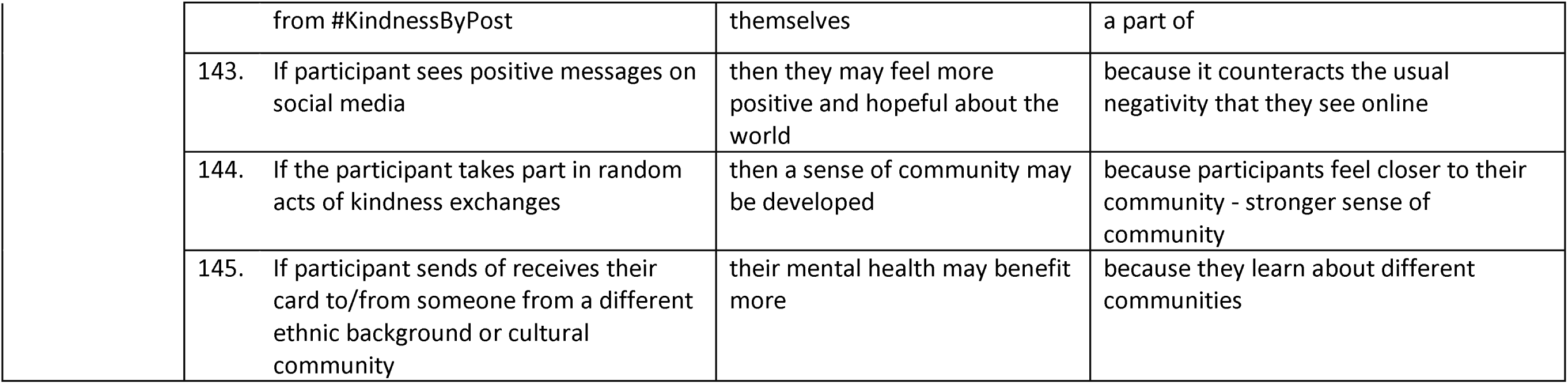

